# Continuous digital collection of patient-reported outcomes during inpatient treatment for affective disorders – implementation and feasibility

**DOI:** 10.1101/2020.08.27.20183400

**Authors:** Maike Richter, Michael Storck, Rogério Blitz, Janik Goltermann, Juliana Seipp, Udo Dannlowski, Bernhard T. Baune, Martin Dugas, Nils Opel

## Abstract

Multivariate predictive models have revealed promising results for the individual prediction of treatment response, relapse risk as well as for the differential diagnosis in affective disorders. Yet, in order to translate personalized predictive modelling from the research context to psychiatric clinical routine, standardized collection of information of sufficient detail and temporal resolution in day-to-day clinical care is needed, based on which machine learning algorithms can be trained. Digital collection of patient-reported outcomes (PROs) is a time- and cost-efficient approach to gain such data throughout the treatment course. However, it remains unclear whether patients with severe affective disorders are willing and able to participate in such efforts, whether the feasibility of such systems might vary depending on individual patient characteristics and if digitally acquired patient-reported outcomes are of sufficient diagnostic validity. To address these questions, we implemented a system for continuous digital collection of patient-reported outcomes via tablet computers throughout inpatient treatment for affective disorders at the Department of Psychiatry at the University of Münster. 364 affective disorder patients were approached, 66.5% of which could be recruited to participate in the study. An average of four assessments were completed during the treatment course, none of the participants dropped out of the study prematurely. 89.3% of participants did not require additional support during data entry. Need of support with tablet handling and slower data entry pace was predicted by older age, whereas depression severity at baseline did not influence these measures. Patient-reported outcomes of depression severity showed high agreement with standardized external assessments by a clinical interviewer. Our results indicate that continuous digital collection of patient-reported outcomes is a feasible, accessible and valid method for longitudinal data collection in psychiatric routine, which will eventually facilitate the identification of individual risk and resilience factors for affective disorders and pave the way towards personalized psychiatric care.

## 1. Introduction

In what has become known as P4 or precision medicine (Hawgood, Hook-Barnard, O’Brien, & Yamamoto, 2015; Lee, Hamideh, & Nebeker, 2019) a major goal of medical research and applied healthcare is the evolution from a reactive treatment approach towards medical care that is predictive, preventative, personalised and participatory.

This approach is of relevance especially in the field of psychiatry, as the imprecise nature of psychiatric nosology, in part due to the heterogeneity of clinical populations, is restricting progress in identifying vulnerable groups and effective treatments (Feczko et al., 2019; Nandi et al., 2009). Affective disorders such as major depressive disorder (MDD) exemplify this problem. Only approximately one third of patients with moderate to severe depression responds to the first treatment attempt with medication (Papakostas & Fava, 2010; Rush et al., 2006). This leads to a prolonged illness duration for non-responders, which is associated with worse overall health outcomes and significantly higher costs to the healthcare system (Ghio, Gotelli, Marcenaro, Amore, & Natta, 2014; Greden et al., 2019). Precision psychiatry could help alleviate this problem by predicting (P1) the occurrence of depression as well as individual disease course and preventing (P2) unfavourable outcomes such as chronification and suicide by personalising (P3) treatment plans according to individual risk and resilience factors.

Preliminary attempts have been made to achieve the prediction of disease course and treatment outcome of MDD through the use of machine learning algorithms (Beijers, Wardenaar, van Loo, & Schoevers, 2019; Cearns et al., 2019; Chekroud et al., 2016; Kessler et al., 2016; Manchia, Pisanu, Squassina, & Carpiniello, 2020). These predictive data-driven approaches are a first step towards the identification of biomarkers for depression, which may ultimately inform clinicians on who is at risk for relapse or a particularly severe outcome and would benefit from more invasive interventions such as electroconvulsive therapy (Redlich et al., 2016). However, previous work relies on extremely homogeneous study populations that are carefully selected according to strict inclusion and exclusion criteria. Any machine learning models trained on such data are of little value when findings are to be generalised to the clinical reality that are highly diverse inpatient populations (Hsin, Fromer, & Califf, 2018; Humphreys & Williams, 2018). In contrast to the aforementioned data from homogeneous, well-characterised study samples, data that is routinely gathered in clinical practice is highly heterogeneous, unvalidated and often not standardized or inaccessible for predictive analysis (De Moor et al., 2015).

In order to achieve the long-term aims of precision medicine in MDD treatment, the implementation of a standardized data collection routine in naturalistic environments in real clinical populations is needed. While high temporal resolution data on sleep and activity levels can be tracked with smartphone or wearable technology based solutions (Bidargaddi et al., 2017), differentiated data on the patients’ mood or affective state – the core features of MDD diagnosis – is needed for precise and valid models of affective disorders that accurately reflect the disease and treatment course. As this information needs to be provided by the patients themselves, much emphasis needs to be put on the participatory (P4) aspect of precision medicine in psychiatry. Patients thus need to be engaged and participate actively by contributing either self-report measures in regular intervals or by participating in clinical interviews or ratings. As external assessments are time-intensive and require clinical training, the use of self-rating scales, which can be completed by patients, independent from the presence of a researcher or clinician might be preferable. Recent evidence suggests reasonably high agreement when comparing patient-reported outcomes (PROs) with diagnostic clinical interviews (Stuart et al., 2014), which supports their use in clinical practice. Previous studies also found the incorporation of PRO into routine care to foster engagement between patients and healthcare professionals and enhance care delivery in various fields of medicine (Lavallee et al., 2016). In psychiatric populations with affective disorders however, questions can be raised as to the patients’ ability and motivation to provide such data, considering the lack of energy as well as cognitive impairments that define MDD during an incapacitating episode requiring inpatient treatment (Cohen, McGarvey, Pinkerton, & Kryzhanivska, 2004; Hindmarch, Hotopf, & Owen, 2013). The difficulty of recruiting depressed patients for randomised controlled trials (RCT) has been well documented (Hughes-Morley, Young, Waheed, Small, & Bower, 2015), although some investigations revealed that patients did report positive attitudes toward research participation when they felt they were contributing meaningfully to the advancement of MDD treatments (Tallon et al., 2011). It remains unclear, how the collection of PROs throughout the treatment course would compare in inclusion rate to the usually much more time-intensive and elaborate study protocols of an RCT. It remains equally unknown whether certain patient subgroups may be systematically less willing or able to provide such data regularly, either due to their symptom severity or other disease-specific or sociodemographic factors, which may constitute exclusion criteria in RCTs.

Another point to consider when striving to make healthcare truly participatory, is that PROs should preferably be collected in a digital format, as digitisation allows for quick data analysis and, ideally, feedback for patients on their personal outcomes (Hirschtritt & Insel, 2018; Hsin et al., 2018). In general, digital solutions outperform paper-pencil questionnaires in practicality, acceptability, and completeness of the data across studies in different fields (Alfonsson, Maathz, & Hursti, 2014; Dale & Hagen, 2007; Fritz, Balhorn, Riek, Breil, & Dugas, 2012). Digital data collection with tablet computers specifically is well-accepted among psychiatric patients (Preuschoff et al., 2013). However, previous investigations with PROs in psychiatry only included a single assessment or pre-post comparisons as opposed to tracking individual outcomes throughout the duration of their hospitalisation (Alfonsson et al., 2014). The feasibility and acceptability of such a study protocol in psychiatric populations remains therefore hitherto unclear.

We established a system of digital continuous data collection, which gives patients the opportunity to participate actively in providing data concerning their mood and symptom levels throughout the course of their inpatient treatment for an affective disorder via tablet computers. This study assessed whether affective disorder inpatients are willing and able to participate in continuous digital data entry throughout the treatment course. We additionally examined whether age, gender, symptom severity and global functioning systematically covary with the feasibility and acceptability of such research efforts in clinical populations with affective disorders. We furthermore aimed to validate patient-reported outcomes of depression severity with the use of an external assessment performed by a clinical interviewer.

## 2. Method

### 2.1 Sample

A total of 364 psychiatric patients that were recently admitted to the inpatient service of the Department of Psychiatry, University of Münster were approached during the assessment period from March 2019 to March 2020. The sole criterion for initial eligibility was inpatient diagnosis of any affective disorder at the time of admission. In order to be included as an active participant, patients needed to be sufficiently mentally stable, cognitively able and proficient in reading and writing German to fill in questionnaires. Due to the naturalistic setting of this investigation, inclusion criteria were intentionally kept as broad as possible in order to achieve the best possible representation of the true population seeking psychiatric inpatient treatment. The study was approved by the local Institutional Review Board (IRB) and written informed consent was obtained before participation. Patients did not receive compensation for their participation.

### 2.2 Procedure

Patients with the appropriate diagnoses were identified with a patient recruitment system (Trinczek et al., 2014) within the electronic health record based on the diagnosis entered by the treating clinician at admission. The clinical team approved research participation for all included patients and could dissent to participation when patients were not suitable due to their mental and cognitive symptom severity or insufficient language skills. All other potential participants were approached in hospital within one week of beginning their inpatient treatment. They were informed about the study and invited to participate for the duration of their stay at several regular intervals. A reason for exclusion was recorded for patients who declined regular participation or were excluded by clinicians.

Upon agreeing to participate, patients were firstly given a tablet-based battery of baseline questionnaires, including questions regarding sociodemographic variables, family and own mental health history, childhood trauma, personality style as well as symptom-specific PROs. External assessments of depressive symptoms and global functioning were additionally conducted by the researcher at baseline. Participants then provided data on their symptom severity in a biweekly rhythm. Immediately before being discharged, they completed selected PROs one more time and were once again assessed externally on their depressive symptoms and global functioning. Please refer to the supplements for more details about the specific measures included in each assessment battery. A researcher was present during data entry, to distribute the tablets and assist patients in case of uncertainty or problems with handling the equipment. The amount of assistance patients required with handling the tablet was rated and the time they took for data entry was recorded immediately after each assessment.

Data was entered via Apple iPads, using the Mobile Patient Survey (MoPat; Soto-Rey et al., 2016), a web-based multi-language electronic PRO system. The standardised data processing and the standardised data export were realised with the single-source metadata architecture transformation (SMA:T; Blitz & Dugas, 2020). SMA:T is an extension of the EHR system and uses Module Driven Software Development to generate standardised applications.

### 2.3 Assessments and measures

Reasons for exclusion were predefined according to the following categories: organisational reasons, severe cognitive deficits, insufficient language skills, and objective mental distress. When eligible participants refused, their reasons for refusal were recorded and later classed into the four categories: lack of interest in the study, subjective mental distress, lack of general compliance, and data security concerns.

An external judgement of patients’ tablet-handling competency was made at each assessment based on a 4-point Likert-scale according to the following categories: 1 – no required support: patient enters data independently; 2 – little required support: patient needs few instructions before entering data; 3 – some required support: patient needs instructions several times during data entry; 4 – a lot of required support: patient largely depends on the researcher for data entry. The median was then calculated from all support ratings.

The researcher kept the time in minutes of each data entry. In order to achieve an individual “entry pace factor”, which signifies the deviation from the group mean, patients’ individual times for the baseline, interim, and discharge assessments were divided by the group mean of each assessment. A mean was once again calculated from these three assessments, resulting in a relational measure of individual data entry pace.

A digital version of the Beck Depression Inventory (BDI; Beck et al., 1996; Hautzinger et al., 1994) was used as a self-report measure of depressive symptoms. The Hamilton Depression Scale (HAMD; Hamilton, 1986) and the Global Assessment of Functioning (GAF; Hall, 1995) were conducted by the researcher as an objective measure of depression severity and global (i.e. psychological, social and occupational) functioning. Please refer to the supplements for an overview of all instruments included in the assessment battery.

### 2.4 Statistical analyses

Statistics were computed using IBM SPSS Version 26.

#### 2.4.1 Participants

##### Required support

To assess the influence of age, gender, depression severity and global functioning on the amount of required researcher support during data entry, we estimated an ordinal logistic regression model that included age, gender, and the baseline sum scores for BDI, HAMD, and GAF as predictors and required support as the dependent variable.

##### Data entry pace

A linear regression model was used to investigate the influence of these same variables on data entry pace. We estimated a linear regression model with age, gender, and the baseline sum scores for BDI, HAMD, and GAF as predictors and entry pace as the dependent variable.

##### PRO validation

In order to validate the self-report measure of depression severity with an external assessment, BDI and HAMD baseline sum scores were firstly correlated. To check for differences in agreement between PRO and external assessments depending on age and gender, we additionally investigated potential interactions with age and gender based on linear regression models. The first model included BDI and age as well as the interaction term age x BDI as predictors and HAMD as the dependent variable. The second model included BDI and gender as well as the interaction term gender x BDI as predictors and HAMD as the dependent variable.

#### 2.4.2 Non-participants

A t-test for independent samples and a chi-square test were calculated to assess whether patients who were excluded by clinicians or study personnel and patients who refused participation differed in age or gender. The same tests were used to assess age and gender differences between participants and non-participants, while potential differences in depression severity and global functioning between these two groups could not be compared, as the data was not available for non-participants.

## 3. Results

### 3.1 Participants

All participants were diagnosed with an affective disorder. The majority of our sample had a diagnosis of MDD (88.8%) and 11.2% had bipolar disorder. 40.1% of the sample were diagnosed with at least one additional psychiatric disorder as a comorbidity, such as anxiety disorders, eating disorders or personality disorders while 16.5% of participants also had a diagnosed somatic comorbidity. On average, participants completed 4 assessments during their hospital stay with a minimum of 1 and a maximum of 15 assessments. No participant dropped out of the study before their scheduled discharge from inpatient treatment.

#### Need for support

A vast majority of participants did not require support and managed data entry independently (89.3%) during all assessments. Little support was needed by 6.6%, whereas 2.9% required some support and 0.8% struggled to enter data independently and relied largely on the researcher for assistance.

For the ordinal logistic regression, predictor variables were tested a priori to rule out violations of the assumption of multicollinearity. Model fit was given (*χ^2^* = 26. 1, *df* = 5, *p* < .001). According to Nagelkerke *R^2^*, the model explained 23.1% of the variance in required support. Age was found to be the only significant contributor to the model as can be seen in **Table 1**. The odds of needing support with data entry and tablet-handling increased with older age (OR = 1.078, 95% CI (1.04, 1.12)). Sex, depressive symptom severity and global level of functioning did not contribute significantly to the model.

**Table 1.** Ordinal logistic regression analysis results predicting the effects of age, sex, and symptom severity on required support during data entry

|  | B (SE) | Wald | OR (95% CI) | *p*-value |
| --- | --- | --- | --- | --- |
| Age | 0.08 (0.02) | 15.96 | 1.08 (1.04 – 1.12) | <.001 |
| Gender (male = 1) | 0.16 (0.54) | 0.09 | 1.18 (0.41 – 3.42) | .76 |
| BDI | -0.03 (0.04) | 0.69 | 0.97 (0.90 – 1.05) | .41 |
| HAMD | -0.08 (0.07) | 1.15 | 0.93 (0.81 – 1.07) | .28 |
| GAF | -0.07 (0.05) | 2.03 | 0.93 (0.85 – 1.03) | .16 |

*Note.* Nagelkerke $R^2 = .231$ .
*Abbreviations.* CI, Confidence Interval; BDI, Beck's Depression Inventory; GAF, Global Assessment of Functioning; HAMD, Hamilton Depression Scale; OR, Odd's ratio; SE, Standard error

#### Entry pace

The linear regression model was significant and explained 31.1% of the variance in data entry time (*R*^2^ = .311, *F*(5,213) = 18.99, *p* < 0.001). Age was a significant predictor of entry pace (*β* = 0.519, *t* = 8.9, *p* < 0.001). Global level of functioning also predicted entry pace (*β* = −0.184, *t* = −2.24, *p* = 0.026) whereas gender and level of depressive symptoms revealed no effect as can be seen in **Table 2**.

**Table 2.** Linear regression results predicting the effects of age, sex, and symptom severity on required time for data entry

| | *B (SE)* | $\beta$ | *T* | *p*-value |
| --- | --- | --- | --- | --- |
| Age | 0.009 (0.001) | 0.519 | 8.899 | <.001 |
| Gender | -0.053 (0.033) | -0.095 | -1.612 | .11 |
| BDI | 0.000 (0.002) | 0.019 | 0.233 | .82 |
| HAMD | -0.001 (0.004) | -0.012 | -0.131 | .90 |
| GAF | -0.006 (0.003) | -0.184 | -2.238 | .03 |

*Note.* $R^2 = .313$ .
*Abbreviations.* BDI, Beck's Depression Inventory; GAF, Global Assessment of Functioning; HAMD, Hamilton Depression Scale; SE, Standard error

#### PRO validation

We found overall high agreement between the patient reported outcome of depression severity and the external clinical rating of depression severity as demonstrated through a strong positive correlation between BDI and HAMD sum scores (*r*(214) = .69, *p* <.001). The additional regression models confirmed these results: The first regression model was significant, with BDI and gender explaining 47.3% of the variance in HAMD (*F*(3,212) = 63.39,*p* < .001, *R^2^* = .473). BDI was a significant predictor (*F*(1,212) = 186.344,*p* < .001) while gender was not. There was no significant interaction between BDI and gender. The second regression model was also significant, with BDI and age explaining 48.2% of the variance in HAMD (*F*(3,212) = 65.863, *p* < .001, R*^2^* = .482). BDI was a significant predictor (*F*(1,212) = 18.191, *p* < .001) while age was not. There was no significant interaction between BDI and age.

### 3.2 Non-participants

Out of the 364 patients that were eligible for inclusion based on their clinical diagnosis, 122 (33.5%) were excluded or refused to participate. The group of non-participants could be split into patients that were excluded by clinicians or study personnel (*n* = 77; 63.1%) and patients that refused participation upon being approached for the study (*n* = 45; 36.9%). There were no differences in age (*t*(120) = 0.207, *p* = .84) or gender (χ^2^(1, *n* = 122) = 0.003; *p* = .96) between these two subgroups.

Half of the patients, who had to be excluded from the study, were excluded due to organisational reasons such as a very short hospital stay (i.e. under one week). Other reasons for exclusions were insufficient German language proficiency, limited cognitive ability (i.e. severe attentional or memory deficits) and acute mental distress as judged by the treating clinician. Within the group of patients, who refused to participate, a majority cited general non-interest in the study as their reason for refusal. Much fewer patients expressed that they felt too incapacitated by their symptoms to participate, displayed general non-compliance with treatment and thusly refused to participate in additional assessments, or expressed concerns over data security. Please refer to **Figure 1** for more detailed visualisation of the distribution of reasons for exclusion and refusal to participate.

**Figure 1.**
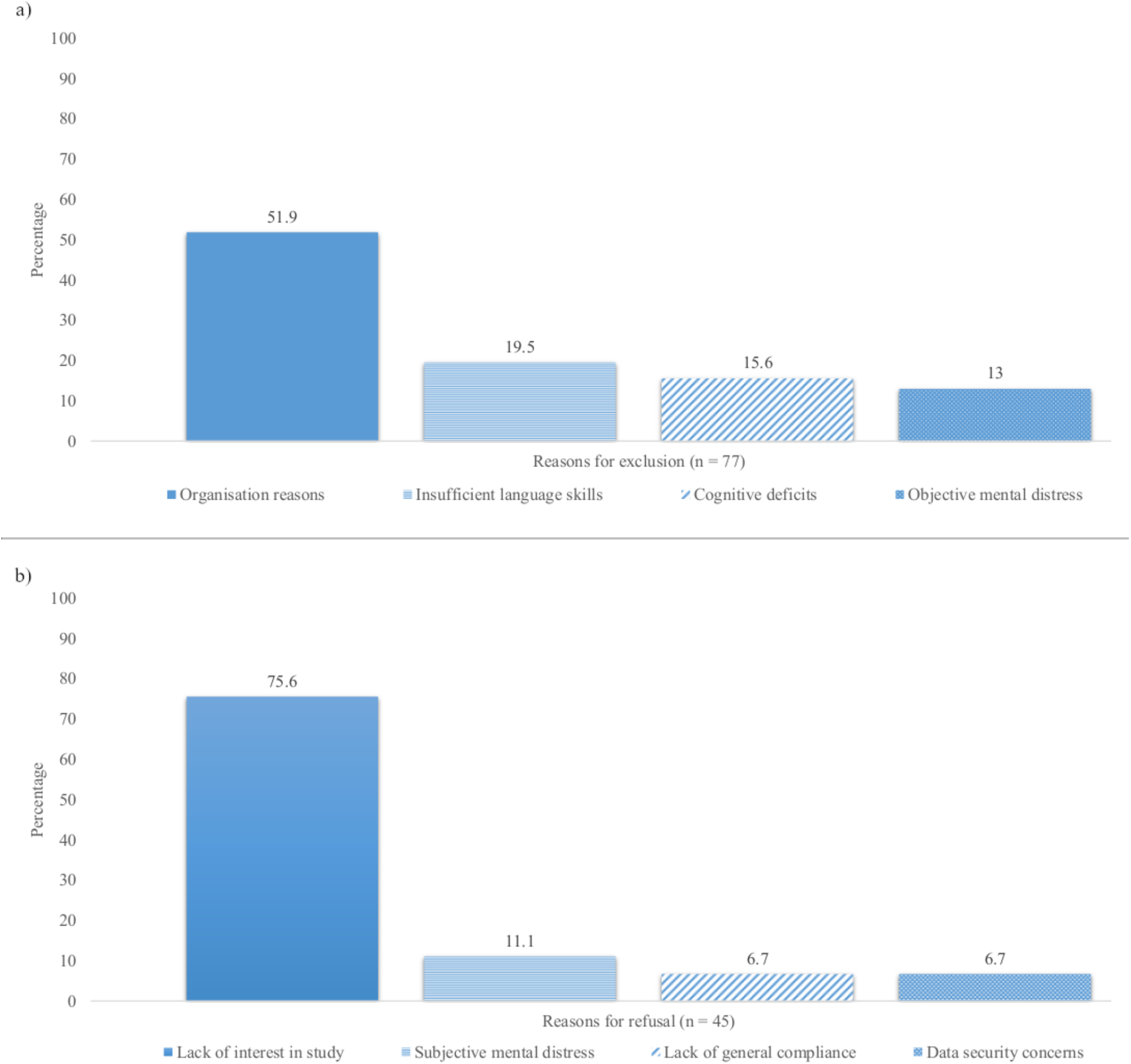
a) Reasons for exclusion of participants as conducted by clinicians or study personnel prior to participation in the study; b) Reasons for refusal of study participation by the patient in percentages from within each non-participating subgroup.

The non-participating group was significantly older (*t*(362) = 3.306, *p* <.001) and consisted of more women (χ^2^(1, *n* = 364) = 4.34; *p* = .04) than the participant group. See **Table 3** for sociodemographic and clinical characteristics of both groups.

**Table 3.** Sociodemographic and clinical characteristics of patients who were initially approached for participation, consisting of 122 non-participating patients and 242 study participants.

|  | Non-participants (*n* = 122) |  |  | Participants (*n* = 242) |  |  | *p*-value |
| --- | --- | --- | --- | --- | --- | --- | --- |
|  | Mean | SD | Range | Mean | SD | Range |  |
| Age | 48.52 | 19.00 | 18 – 89 | 41.93 | 17.39 | 18 – 81 | < .001 |
| Gender (m/f) | 43/79 |  |  | 113/129 |  |  | .04 |
| BDI | N/A |  |  | 24.09 | 11.24 | 1 – 50 |  |
| HAMD | N/A |  |  | 15.53 | 6.16 | 1 – 31 |  |
| GAF | N/A |  |  | 57.44 | 8.63 | 33 – 78 |  |

*Abbreviations.* BDI, Beck's Depression Inventory; GAF, Global Assessment of Functioning; HAMD, Hamilton Depression Scale; SD, Standard deviation. Means, standard deviations (SD) and group differences (as measured with t-test or $\chi^2$ -test).

## 4. Discussion

With this study, we demonstrate the feasibility and acceptance of digital continuous data collection in affective disorder inpatients. Our results indicate that participatory medicine can be achieved in patients with affective disorders, as they are willing and able to contribute PROs throughout the duration of their inpatient treatment. This study therefore provides important insight into the possibility of routinely collecting longitudinal data in real-world clinical cohorts that may guide the way towards personalised psychiatric care.

During the assessment period of one year, we achieved an inclusion rate of 66.5% of patients with a diagnosed affective disorder. This rate is similar to that reported from other investigations performed on the general population and non-psychiatric patient groups, which indicates that MDD symptomology does not constitute a barrier towards participation (Grobbee et al., 2005; Karsten et al., 2018). Exclusion by the clinician or researcher and refusal to participate by the patients themselves were largely not due to symptom severity or cognitive impairment but for organisational reasons or a general disinterest in the study, which mirrors reasons for non-participation in research from non-psychiatric populations (Brintnall-Karabelas et al., 2011).

A vast majority of patients who did participate were able to enter data independently and did not encounter technical difficulties. More importantly, we found no association between symptom severity at baseline and the amount of required support in handling the equipment or prolonged data entry times. This is in line with previous investigations on the feasibility and acceptability of digitally based assessments in psychiatric populations (Preuschoff et al., 2013). Moreover, we were able to demonstrate the validity of PROs with the use of an external measure of depression severity performed by a clinical interviewer. The level of agreement between self-reported depression severity via the BDI and the external rating based on the HAMD was comparable to findings from the literature (Steer, Beck, Riskind, & Brown, 1987) and indicated high validity of the digital PRO. This is an especially promising result for the implementation of PRO collection technologies into routine documentation as well as its use for research purposes as it indicates that little to no additional personnel resources are required in order to gain valuable longitudinal data throughout the course of treatment. Moreover, the digital implementation of PROs will allow for more precise and immediate data storage and analysis (Hsin et al., 2018). In the future, such an infrastructure of digital data collection could be used to communicate treatment outcomes and visual representations thereof directly to patients. Similar approaches have been found to improve communication between patients and healthcare providers (Baeksted et al., 2017), and would also constitute an improvement in the participatory aspect of precision medicine.

Although our results generally support the feasibility of continuous digital data collection in affective disorders, a few systematic recruitment and accessibility issues must be addressed. Despite the fact that no association between symptom severity and performance during data entry in our participating sample was detected, 6% of participants were excluded beforehand due to reduced cognitive ability or clinicians’ concerns over their acute mental distress. Although this embodies only a small percentage of our sample, this result suggests that a systematic exclusion of more severe cases may not be avoidable. Nevertheless, it may be worthwhile to critically consider such cases individually, as it has been shown that carers overestimate the amount of distress patients are put under during research participation (Tallon et al., 2011).

We furthermore found that women were more likely to refuse or be excluded from the study than men. This may again be due to symptom severity. Although the reasons recorded at the time of exclusion do not suggest symptom severity to be the main factor, evidence does suggest women to be more likely to seek psychiatric treatment and report more severe symptoms (Koopmans & Lamers, 2007; Ladwig, Marten-Mittag, Formanek, & Dammann, 2000). As we do not have data on the symptom levels of the excluded group, this question cannot be answered with certainty.

Regardless of gender, the factor that impacted both the amount of required assistance and time during data entry was older age. Older adults found it more difficult to handle the tablet-based assessments and took longer to complete them. However, the percentage of participants who needed assistance was comparatively small and even those who did require assistance were able to complete assessments regularly, which suggests that their difficulties with handling the equipment did not stop them from participating. Although our study did not assess subjective attitudes toward technology, previous studies found that digital methods of data collection are well-accepted even among older adults (Horevoorts, Vissers, Mols, Melissa, & Van De Poll-Franse, 2015; Karsten et al., 2018). It can also be expected that technological literacy will rise in older populations over the years, as smartphones and tablets are becoming increasingly ubiquitous, which will alleviate the difficulties for this specific age group in the future. Studies also show that, although older adults lag behind in digital literacy, such competencies can be acquired through social support (Schreurs, Quan-Haase, & Martin, 2017; Tsai, Shillair, & Cotten, 2017). Nevertheless, our findings suggest, that options for support of older participants should currently be offered so as not to systematically exclude technologically less well-versed patients from participatory care.

In addition to older age, lower global functioning was also related to slower data entry pace; however, it was not associated with the amount of required assistance. This suggests that patients with a generally lower level of global functioning take longer to complete assessments but are still able to do so independently. Moreover, the added time expenditure does not lead to participants dropping out of the study, indicating that the slower entry pace is tolerable and not a barrier that would keep lower functioning patients from participating in such research.

Lastly, those patients who are not proficient in the language spoken by their healthcare providers are a systematically disadvantaged group in psychiatric care that could not be included in this investigation. At equal or greater levels of need, migrants are known to seek mental health treatment less often and are less likely to report favourable treatment outcomes (Derr, 2016; Mösko, Schneider, Koch, & Schulz, 2008). This suggests that the inclusion of marginalised populations would be of great importance especially when investigating individual risk factors for affective disorders on the way to precision psychiatry. In fact, digitally assessed PROs present the opportunity to get detailed, standardised assessments despite language barriers, as questionnaire measures can be made available in every language. The application we used for data collection supports the implementation of multi-language assessments (Soto-Rey et al., 2018) and could therefore be used in future investigations in order to also reach and assess non-German speaking clinical populations. This would provide a wealth of standardised, quantifiable information about patients of diverse cultural backgrounds that could guide treatment but also assist in identifying suitable interventions for clinical populations with specific ethnic or cultural differences and risk or resilience factors.

To the best of our knowledge, this study is the first naturalistic investigation to incorporate continuous, digitally assessed PROs throughout the course of inpatient treatment in affective disorders. Overall, the acceptability and feasibility of such study protocols within the clinical routine is high while required resources remain comparatively low. Patients are willing and able to provide data in regular intervals and are not systematically disadvantaged by the severity of their affective symptoms. Future implementations should keep gender, age, and cultural factors in mind when approaching patients and offer assistance with any technological equipment as needed.

In conclusion, this study is a first step in demonstrating that the participatory aspect of precision medicine can be achieved in psychiatry. In the future, the information gathered routinely through patient reported outcomes may be combined with other potential data sources such as fitness trackers and information gained from electronic health records (Goldstein-Piekarski, Holt-Gosselin, O’Hora, & Williams, 2020; Hariman, Ventriglio, & Bhugra, 2019; Hirschtritt & Insel, 2018; Jensen et al., 2015). This will pave the way for data-driven machine learning models that could ultimately be used to predict and prevent the occurrence of affective disorders, as well as facilitate the identification of individual risk profiles. Such advances in psychiatry will be invaluable as personalised treatments tailored to such individual risk factors may lead to much shorter and less frequent hospitalisations, which would equate to more cost-effective treatments and a pronounced reduction in patient suffering.

## Data Availability

Derived data supporting the findings of this study are available from the corresponding author on request.

## Funding and disclosure

Funding was provided by the Interdisciplinary Center for Clinical Research (IZKF) of the medical faculty of Münster (Grant SEED 11/19 to NO), as well as the “Innovative Medizinische Forschung” (IMF) of the medical faculty of Münster (Grants OP121710 to NO).

## Acknowledgements

We are deeply indebted to all participants of this study.

## Notes

### Competing Interest Statement

The authors have declared no competing interest.

### Funding Statement

Funding was provided by the Interdisciplinary Center for Clinical Research (IZKF) of the medical faculty of Muenster (Grant SEED 11/19 to NO), as well as the Innovative Medizinische Forschung (IMF) of the medical faculty of Muenster (Grants OP121710 to NO).

### Author Declarations

Ethik-Kommission der Aerztekammer Westfalen-Lippe und der Westfaelischen Wilhelms-Universitaet, Muenster, Germany

